# Spectrum of HRCT chest findings in RT-PCR positive asymptomatic COVID-19 patients at a COVID designated hospital in Nepal

**DOI:** 10.1101/2021.06.25.21259523

**Authors:** Bina Basnet, Sujit Pant, Sujata Pant, Kalpana Rai, Niraj Basanta Tulachan, Bibek Karki, Rajiv Shahi, Shiva Bahadur Basnet, Bikash Bikram Thapa

## Abstract

**Introduction:** COVID-19 pandemic is grappling the world with the surge of infection time and again. Clinicians are trying to justify the ethics of public health care. Asymptomatic COVID-19 cases are going undocumented and most of them practice self-isolation. Studies have revealed significant radiological changes among RT-PCR positive asymptomatic COVID-19 cases.

**Objective:** The aim of this cross-sectional study is to characterized chest CT findings of asymptomatic RT-PCR-positive patients in one of the COVID-designated hospitals in Nepal.

**Results:** Out of 43, 26 (60.5%) participants had positive Chest CT scan findings consistent with COVID pneumonia. 65% had bilateral and 77% had multifocal lesions. The ground-glass opacities (92%), mixed (ground-glass opacities and consolidation) pattern (30.7%), and consolidation only (34.6%) were common chest CT findings. The median CT score was 3.5 (Interquartile range; 2-6).

**Conclusion:** The majority of the RT-PCR positive asymptomatic patient present with CT scan changes of lungs which are important to determine clinical status, prognosis, and long-term sequel in those cohorts.

## Introduction

Since the first reports of the Corona Virus Disease-2019 (COVID-19) from Wuhan, city in China, the cases have been reported from all the seven continents and had already caused death in millions. The COVID-19 disease caused by severe acute respiratory syndrome coronavirus 2 (SARS-CoV-2) is primarily transmitted through respiratory droplets and infect lungs. [1] Reverse Transcriptase Polymerase reaction (RT-PCR) is the confirmatory laboratory diagnosing test for SARS-CoV2. The RT-PCR is recommended in asymptomatic cases especially in close contacts, screening and early identification of infection in high risk population and locations like healthcare centers, prior to surgical procedures, and prior to receiving immunosuppressive therapy. [2,3]

In a metanalysis the pooled percentage of the asymptomatic infection was 15.6%. [4] Asymptomatic cases are less infectious than symptomatic. [5,6] Despite low infectivity, 20-50% of the asymptomatic cases had specific changes in the chest computed tomography (CT). But the probability of progression of the asymptomatic cases into pneumonia is very less (<1%). [7,8] CT changes of the asymptomatic patients are important not only to evaluate the chest CT as alternative diagnostic modalities for selected COVID-19 cases but also to evaluate the long term outcomes of COVID-19 related respiratory pathophysiology, which is mostly unknown. [9]

The objective of our study is to characterize the chest CT scan features and clinical outcomes of RT-PCR confirmed asymptomatic COVID-19 patients in COVID-19 designated hospital (Shree Birendra Hospital), Nepal.

## Methodology

This is a cross sectional study where a cohort of laboratory proven RT-PCR positive COVID-19 asymptomatic adult (age > 18 years) patients were included and were subjected to the HRCT scan of the chest. All the cases included in this study fulfill the WHO criteria of the “close contacts” of the COVID-19 confirmed cases. The study is conducted between October 2020 and December 2020 with ethical approval from institutional review board and consent of the patient and

HRCT scan were acquired in the Hitachi Multidetector 128 slice CT scanner. The parameters for used for CT acquisition were helical mode volumetric HRCT with Tube voltage 100kVp-120kVp and tube current 80-500mA, and slice thickness of 1.0mm with reconstruction interval, 0.6mm using a sharp reconstruction algorithm. CT images were obtained with the patient in supine position with full inspiration. Intravenous contrast administration was not used. Acquired images were transferred to a separate workstation for further processing. Image was reconstructed in axial, coronal, and sagittal planes to detect the craniocaudal and axial/peripheral distribution of the lung parenchymal involvement.

All images were viewed on both lung (width, 1500 HU; level, −700 HU) and mediastinal (width, 350 HU; level, 40 HU) settings. The chest CT scan was evaluated by two radiologist characterizing the parenchymal involvement on the basis of the (a) characteristic findings and morphology: ground glass opacities, consolidation, linear bands, bronchial wall thickening, nodules and additional findings like pleural effusion and mediastinal lymphadenopathy. (b) Distribution of the involvement: Laterality, craniocaudally distribution, number of the lobes involved, percentage of involvement in each lobe. Then the CT severity score (0-25) was calculated following the semi-quantitative scoring system which depends on the visual assessment of the each 5 lung lobes (0-0%; -<5%; 2-5 to 25%; 3-26 to 50%; 4-51 to 75%; 5-> 75%) that was initially proposed by Pan et al.[10]

## Results

In this cross-sectional study total of 43 RT-PCR positive COVID-19 cases were included. Among the study cohort 26 (60.5%) had positive Chest CT scan findings (Figure 1-3). The positive Chest CT had mostly bilateral (65.3%) and multifocal (77%) lesion. 92% of them had ground glass opacities (Table 1).

**Figure 1:**
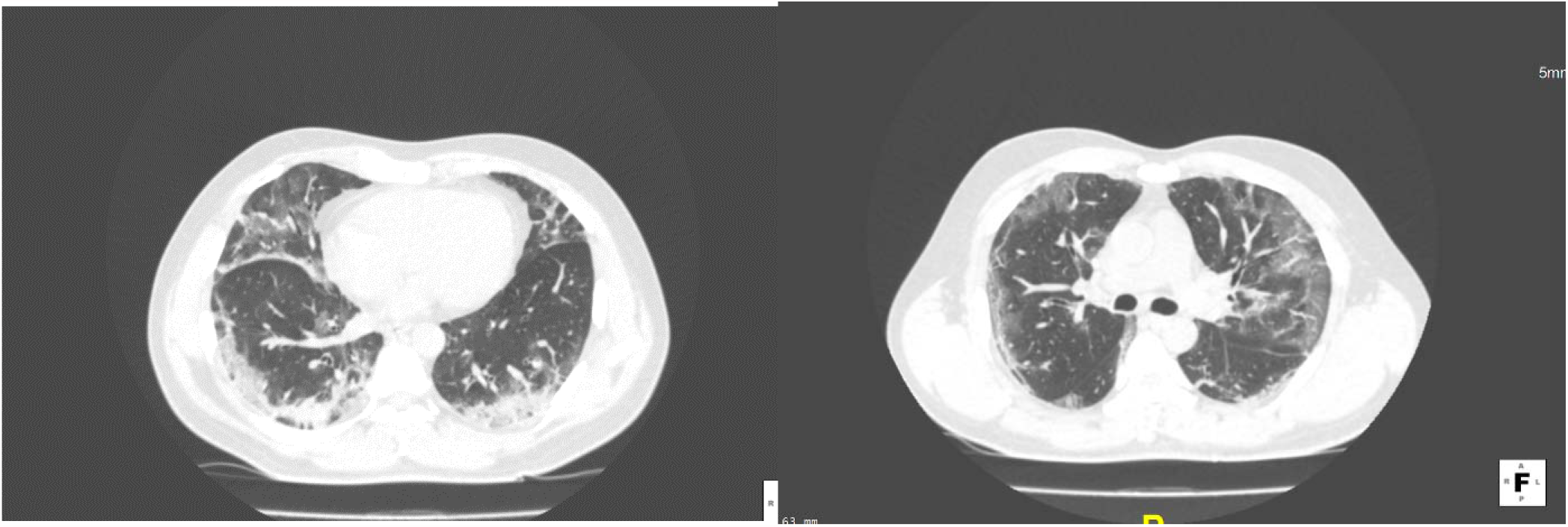
Multifocal predominantly peripheral consolidative and ground glass opacities in bilateral lung. Interspersed areas of interlobular septal thickening seen in the inferior lingual segment of left upper lobe.

**Figure 2:**
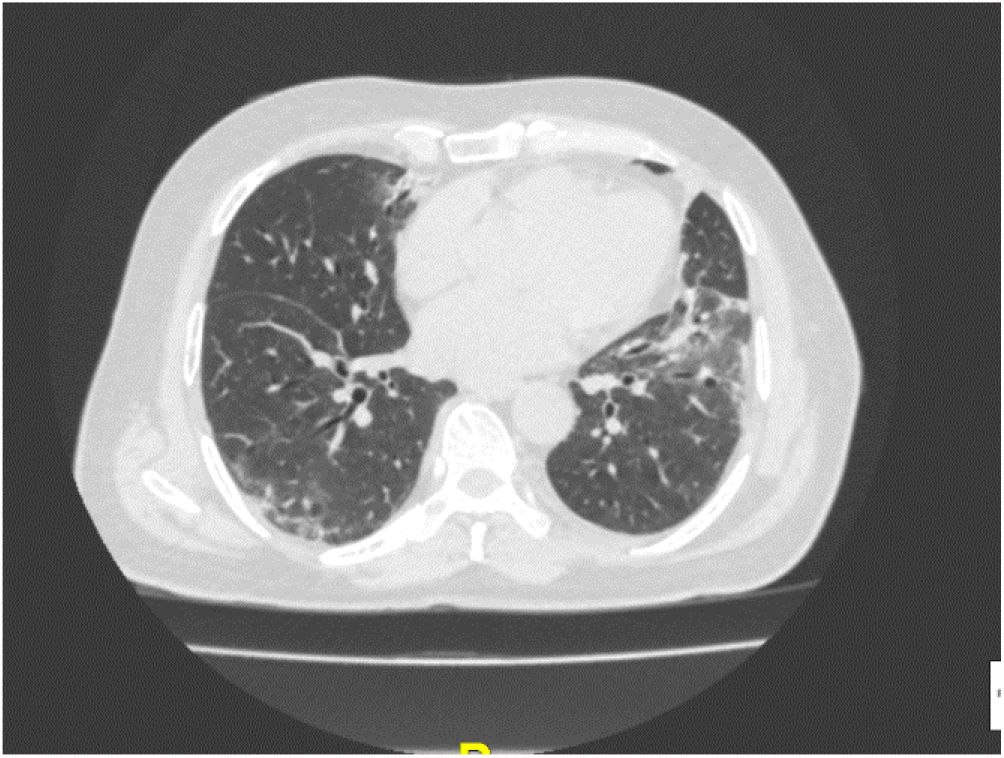
Patchy peripheral ground glass opacity with focal traction bronchiectasis within the affected area

**Figure 3:**
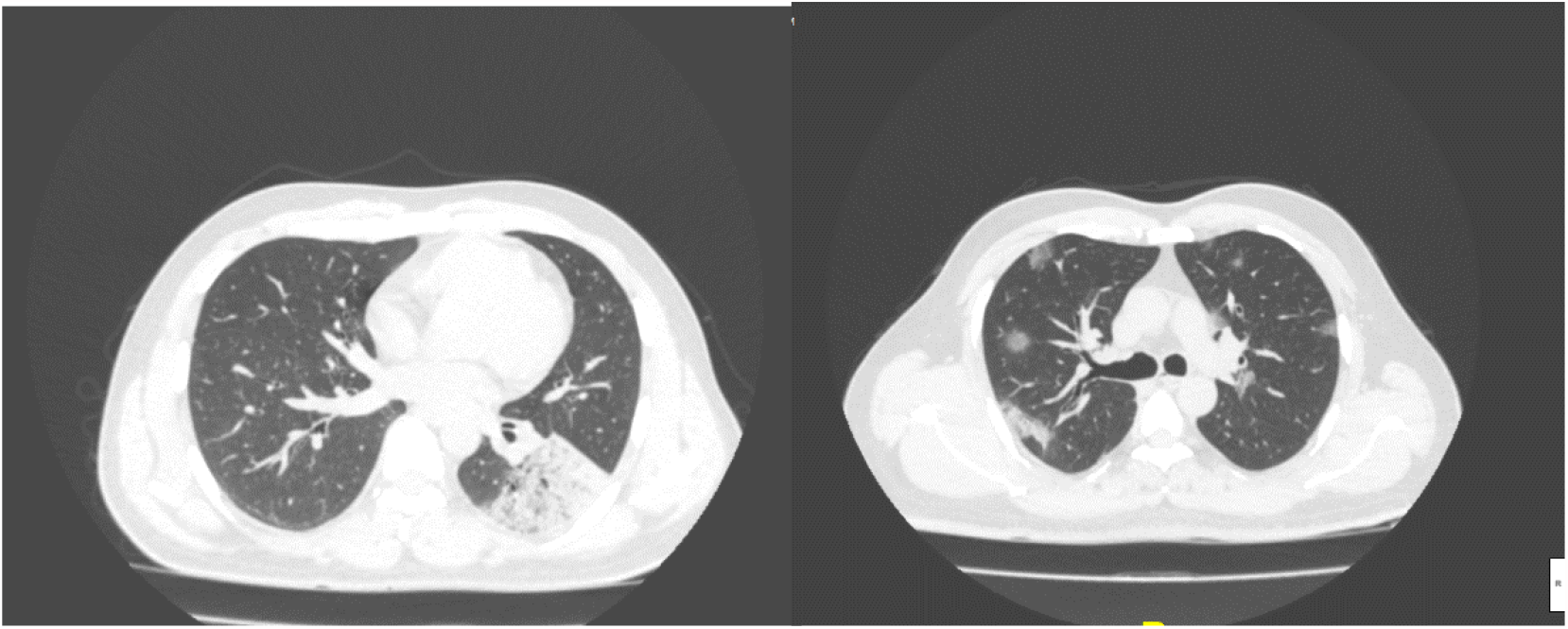
Consolidation (a) Subsegmental (b) multifocal peripheral rounded morphology

**Table1:** HRCT details of the study cohort:

| Particulars | Value |
| --- | --- |
| Age | 41.42±11.7 (23-66) |
| Male: Female | 3:2 |
| Positive CT findings | 60.5%(26/43) |
| Lobar involvement | Right- 7 (27%) , Left-2 (8%) and Bilateral -17 (65%) |
| Number of Focus | Single- 23% (6/26; Multiple – 77%(20/26) |
| Characteristic Chest HRCT scan findings | Consolidation 34.6%; (9 )<br>Ground Glass opacities 92%; (24)<br>GGO and Consolidation 30.7% (8)<br>Atelectatic Band 19% (5)<br>Septal thickening 30.7%; (8) |

The median global CT score of the abnormal CT scan chest was 3.5 (Interquartile range; 2-6). The mean CT score value was significantly higher (5.3±2.6 vs 3.0± 1.0) in bilateral lung disease. The ratio of upper to lower lobe involvement was 2:3. The patients with normal CT scan had higher mean cycle threshold (CT) value of RT-PCR test than abnormal chest CT scan group (23.8 vs 21.7;). However, the difference was statistically not significant. One patient developed mild COVID-19 symptoms (cough and headache) during follow up.

## Discussion

Pneumonia is primary manifestation of the COVID-19 disease. WHO has classified symptomatic COVID-19 infection into mild cases, pneumonia, severe pneumonia, and critical disease (sepsis, septic shock and or ARDS). In a large cohort of population based study the 30-40% were asymptomatic young adults. The virus can be cultured from infected individual as early as six days prior to the developments of symptoms. Whereas it takes at leas a week for CT scan to detect changes in lung parenchyma.[11-14] Screening CT chest had positivity rate of 1.6%.[15] In RT-PCR confirmed COVID-19 cases HRCT help in prognostication, evaluating the disease progression and monitoring the response to therapy. [16]

With RT-PCR as reference, the sensitivity, specificity, accuracy of chest CT in indicating COVID-19 infection were 97%, 25% and 68% respectively, the accuracy of which is higher in age more than 60 years. Studies have shown that Bilateral lung (90%) involvement with ground glass opacities (46-50%) and mixed GOO along with consolidation (44-50%), and consolidation only (25%) were the most common pattern in Chest CT scan characteristics of COVID-19 patients. [12,14,16,17,16] Similar pattern of Ground glass opacities (95%) and consolidation (5%) with predominantly bilateral, sub pleural, and multiple lesion were described among asymptomatic cases.[9] [18] 83% had features of lung consolidation among asymptomatic COVID-19 cases (54%) found in the cruise ship “Diamond Princess”.[19] Parenchymal inflammation, endothelial dysfunction, and cytokine release syndrome are responsible for COVID-lung and final radiological manifestations. The pathophysiological pathway can lead to one or more of the following: acute respiratory distress syndrome (ARDS) with diffuse alveolar damage (DAD), diffuse thrombotic alveolar microvascular occlusion, and inflammatory mediator-associated airway inflammation.[17,20,21,20]

The median CT severity score in this study was 3.5 with range (range, 1-11). Francone et all found significant positive correlation (p< 0.0001) of CT score between age of patient, inflammatory biomarkers, and with severity of the disease. CT score >18 has hazard ratio of 8.33 (95% CI, 3.19-21.7) for COVID-19 related mortality.[22] The significance of CT score in asymptomatic patients is yet to be evaluated in short and long term follow up.

Based on SARS-COV-1 data of 2003 two third of the survivors suffered from TGF-β-mediated pulmonary fibrosis and SARS-COV-2 is expected to share similar chronic sequel.[23,24]. CT chest is widely available modality to assess and follow the pulmonary changes in COVID-19 patients both symptomatic and asymptomatic. During early 2020 asymptomatic or undocumented cases were responsible for 79% of the documented cases. [25]Probability of having incidental CT findings among asymptomatic cases is high due to ongoing progression of the pandemic. Though it is difficult to explain the temporal phase of CT changes in asymptomatic cases screening CT is recommended for all RT-PCR positive COVID-19 patients for the purpose of characterization of the findings and its long-term sequel.[26] The study was conducted during first COVID-19 pandemic wave in Nepal when the average nation wide case positivity rate was 15.6%. Study with large sample size and follow up scan add more to the scientific evidence on radiological features of asymptomatic COVID-19 cases. Study in a context of higher incidence rate can unfold different data and evidences.

## Conclusion

Chest CT plays an important role in the diagnosis and management of the RT-PCR negative COVID suspected as well as RT-PCR positive asymptomatic COVID-19 cases. The long term sequel of the COVID-Lung is little known to us. The present evidences suggest that the COVID-19 asymptomatic cases should be followed clinically and radiologically in order to evaluate its outcome.

## Data Availability

All data is available for review by contacting Dr. Bikash Thapa.

## Declarations

### Ethics approval

Ethical approval for this study was taken from Institutional review board

### Consent to participate

Written consent taken

### Consent for publication

All authors consent to publication of this paper

### Availability of data and material

All data is available for review by contacting Dr. Bikash Thapa

### Code availability

Not applicable

### Authors’ contributions

All author has made substantial contributions to the conception or design of the work; the acquisition, analysis, interpretation of data; and the creation of manuscript. BB agreed to be accountable for all aspects of the work in ensuring that questions related to the accuracy or integrity of any part of the work are appropriately investigated and resolved.

